# Accumulation of immunity in heavy-tailed sexual contact networks shapes monkeypox outbreak sizes

**DOI:** 10.1101/2022.11.14.22282286

**Authors:** Hiroaki Murayama, Carl A. B. Pearson, Sam Abbott, Fuminari Miura, Sung-mok Jung, Elizabeth Fearon, Sebastian Funk, Akira Endo

## Abstract

Many countries affected by the global outbreak of monkeypox in 2022 have observed a decline in cases. Our mathematical model incorporating empirical estimates of the heavy-tailed sexual partnership distribution among men who have sex with men (MSM) suggests that monkeypox epidemics can hit the infection-derived herd immunity threshold and begin to decline with less than 1% of sexually active MSM population infected regardless of interventions or behavioural changes. Consistently, we found that many countries and US states experienced an epidemic peak with cumulative cases of around 0.1–0.7% of MSM population. The observed decline in cases may not necessarily be attributable to interventions or behavioural changes primarily, although continuing these approaches in the most effective manner is still warranted to minimise total epidemic size.

## Main text

Since May 2022, sustained local transmission of monkeypox has been confirmed in Europe, the Americas and other regions where the virus was not observed to circulate previously. Prior to this global outbreak, monkeypox was considered to be primarily driven by exposure to animal reservoirs with limited transmission potential among humans (*1, 2*). Yet, in the 2022 outbreak, observed cases have been predominantly among men who have sex with men (MSM) with no reported exposure to animals or travel history in endemic countries and cases among other population groups have been limited (*3*). This novel outbreak profile can be explained by sexually-associated transmission (*4*) and a heavy-tailed empirical distribution of sexual partners among MSM, which could lead to sustained human-to-human transmission in this population while not in others (*5*). Monkeypox is known to be transmitted through skin-to-skin contact, droplets and fomites (*6*). In previous monkeypox outbreaks, studies estimated the secondary attack risk (SAR) in unvaccinated household contacts to be around 10%, though did not evaluate particular risk to sexual partners (*7*). The SAR specifically among sexual partners remains an open question, but a wide range of sexual SAR values would lead to sustained outbreaks over MSM sexual contact networks (*5*).

The rapid initial surge of cases with male predominance reported across affected countries has been consistent with this theory; however, as of November 2022, many of those countries have seen an apparent slowdown in growth of cases followed by a decline. This shift in trends may be contributed to by various reactions since the identification of the current monkeypox outbreak, including public health interventions such as contact tracing and vaccination (*8*–*10*) and heightened awareness triggering behavioural changes among high-risk populations (*11*). However, available evidence is overall insufficient to quantify the relative contribution of these responses to the decline in different countries and operational indicators suggest impact may have been blunted by practical factors. Contact tracing and ring vaccination have, at times, been faced with untraceable contacts and limited consent rate (*9, 12*). Vaccine supplies were initially limited, slowing rollout of mass vaccination and precluding many countries from achieving substantial coverage before observing a peak (*12, 13*)—moreover, time required for eligible individuals to complete the dosing schedule (e.g. 2 doses 4 weeks apart for JYNNEOS vaccine in the US (*14*)) and for immunity to be established (suggested to be up to two weeks by public authorities (*15*) although evidence remains limited (*16*)) renders prompt epidemic control by vaccination more challenging. Providing a coherent explanation to the observed decline in growth in many affected countries at different times and outbreak sizes is not straightforward.

Another key mechanism that can shape epidemic trends is accumulation of infection-derived immunity, known as (infection-derived) ‘depletion of susceptibles’ or ‘herd immunity’ (*17*). Highly heterogeneous contact patterns are known to lead to a high basic reproduction number (*R*_0_) but lower the herd immunity threshold for immunising infections (*18*–*21*)—i.e. when a small fraction of individuals exhibit disproportionately high contact rates, the initial epidemic growth could be accelerated by transmission among these individuals but this growth would also be short-lived as these individuals become rapidly infected and immune and no longer contribute to the outbreak. The heavy-tailed nature of the sexual partnership distribution among MSM could create these conditions and thus explain the initial growth of monkeypox cases in many affected countries (*5*) but also their quick saturation. Without accounting for such inherent saturation effects, analysis of monkeypox case trends may incorrectly attribute declines to other factors. To better understand the current dynamics of monkeypox, we need to illustrate the baseline epidemic trajectory anticipated under the sole effect of infection-derived immunity, absent any responsive changes to transmission patterns.

We developed a mathematical model of monkeypox transmission over the MSM sexual contact network that accounts for infection-derived immunity. Our model suggested that, with a plausible SAR in a highly heterogeneous sexual contact network consistent with the observed heavy-tailed sexual partnership distribution among MSM, an epidemic rapidly hits the herd immunity threshold and starts to decline. This may explain the current decline in monkeypox cases in many countries with diverse timing and intensity of interventions. We found that many of the observed monkeypox epidemics formed a peak when the cumulative number of cases reached about 0.1–0.7% of estimated sexually active MSM population size—such patterns are reproduced by our model with a SAR of between 10–30% per sexually-associated contact without assuming any impact of interventions or behavioural change.

In our model, we represented the heavy-tailed distribution of sexual partners among MSM over the infectious period of monkeypox (assumed to be 14 days) as a left-truncated Weibull distribution parameterised in our previous study (*5*) using the British National Survey of Sexual Attitudes and Lifestyles (Natsal) data (*22*). We assumed that non-MSM transmission dynamics is negligible because transmission over MSM sexual networks could well approximate the overall dynamics of monkeypox in the current outbreak given their predominance among cases (> 95%) (*3*). The risk of an individual being in contact with an infectious sexual partner was modelled as proportional to the number of their sexual partners over 14 days. Upon recovery, infected individuals were assumed to develop long-term immunity and maintain their sexual behaviour without further risk of re-infection. To improve robustness of the model to uncertainties in time-related parameters such as generation time and reporting delay, we used cumulative incidence as a measure of epidemic progression instead of time—i.e. we directly modelled the relationship between the cumulative number of cases per MSM population and the effective reproduction number *R*_eff_.

To compare our model outputs with observed monkeypox outbreak data, we identified the period during which reported cases likely peaked in different populations (European countries, the US, Canada and US states). We fitted Gompertz curves to the cumulative reported case count over time in each of the included countries and US states and estimated the cumulative number of monkeypox cases per MSM population size (*23, 24*) by the apparent epidemic peak (cumulative incidence proportion at a peak of an epidemic; CIPP), where the estimated daily epidemic growth rate is consistent with a near-zero value (i.e. within ±0.01). We defined the “consensus range” as a set of values that lies within the CIPPs of at least 50% of included countries/states. That is, any value within the consensus range is consistent with the majority of the country/state CIPPs. The consensus range among the included countries suggested that their epicurves were generally consistent (though with some apparent outliers) with a saturation of growth when the cumulative case count reached 0.13%–0.39% of the estimated MSM population size (Fig. 1A). Moreover, 17 out of 24 (71%) countries had their CIPP ranges overlapping at 0.24–0.27% (Fig. S5A). The consensus range among US states was 0.14%–0.65% and CIPPs of 31 out of 45 (69%) states shared 0.21–0.26% in common (Fig. 1B, Fig. S5B). We did not find a clear correlation between CIPP and the number of allocated vaccine doses per MSM population by the peak among US states (Spearman’s correlation 0.21 [95% confidence interval -0.12, 0.53]) (Fig. S1) but found that states with later epidemic onset (defined as the date of reporting the 10th case) tend to have lower CIPPs (correlation -0.65 [-0.84, -0.37]) (Fig S2). We also found a similar but non-significant correlation between CIPP and epidemic onset among the included countries (correlation -0.60 [-0.91, 0.11]).

**Fig. 1.**
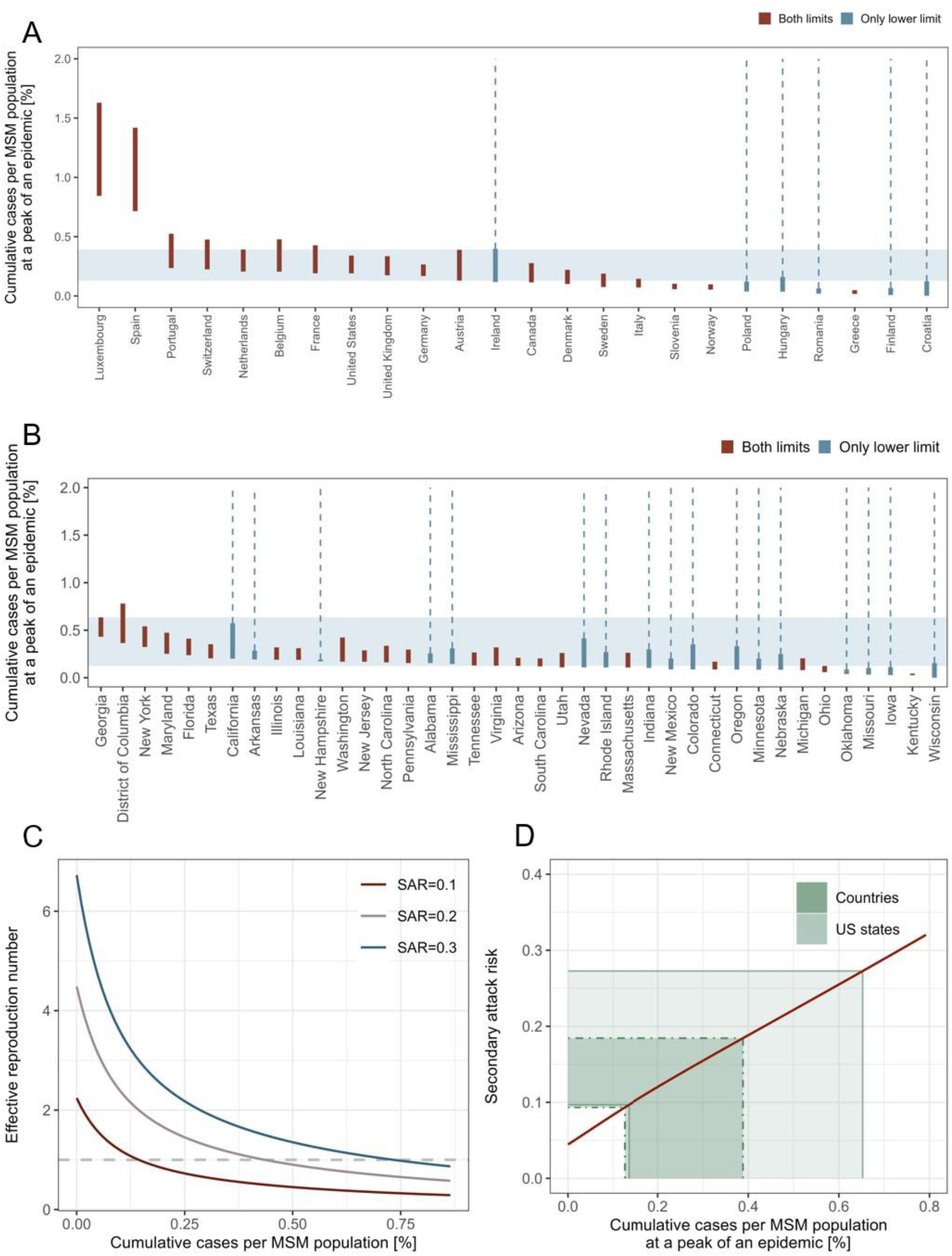
The observed and modelled number of cumulative monkeypox cases per MSM population. (A, B) Estimated range of cumulative incidence proportion at the peak of an epidemic (CIPP) (A) by country and (B) by US state. Some countries or US states have not clearly passed the peak as of available data (last updated on 15 October 2022) and therefore the upper limit of CIPP is undetermined (blue bars); others have apparently passed the peak and have both limits for CIPP (red bars). The consensus range of CIPP (values consistent with at least 50% of included countries/states) is shown with light blue shades. (C) Modelled trajectory of the effective reproduction number (R_eff_) over the course of an epidemic. The reproduction number was computed for three possible values of SAR (0.1, 0.2, and 0.3). (D) Estimated relationship between CIPP and SAR. Thick and thin green areas represent the global and US consensus ranges of CIPPs, respectively.

Our model of accumulation of infection-derived immunity in a heavy-tailed MSM sexual contact network can explain these epidemic peak sizes falling in a similar order of magnitude. Since our model is scale-invariant, in the absence of exogenous influences such as interventions and behavioural changes, we anticipate identical CIPPs across different MSM populations if they share the same partnership distribution and SAR. As individuals with highest numbers of partners are most likely to be infected in the earliest phase of an epidemic, the effective reproduction number *R*_eff_ would rapidly decline in this case as transmission progresses. Considering SAR values of 10, 20 and 30%, our model found that, while *R*_0_ (the initial value of *R*_eff_) is well above 1, *R*_eff_ rapidly decreases and crosses 1 after observing relatively few cases (< 1% of the MSM population) (Fig. 1C). The herd immunity thresholds given an SAR of 10%, 20% and 30% were estimated to be 0.15%, 0.43%, and 0.74% of the MSM population, respectively. These thresholds are substantially lower than the classical herd immunity threshold in a homogeneous population: 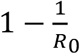 (55%, 78% and 85%, respectively, based on the values of *R*_0_ in our model). We showed in Fig. 1D that the observed consensus ranges of CIPPs are consistent with SARs of around 10%–20% (global) or 10%–30% (US states) if they are formed primarily by infection-derived immunity and our model assumptions are valid. We also estimated the final size of an epidemic driven by infection-derived immunity alone corresponding to different SAR values (Fig. 2). The estimated final epidemic size was generally more than double the size of CIPP, contrary to outcomes for a conventional homogeneously mixing transmission model (see *Supplementary Materials*). This suggests that the decreasing phase of an epidemic with highly heterogeneous transmission patterns may be more gradual than that of a homogeneous epidemic. The estimated final size relative to CIPP increased with SAR.

**Fig. 2.**
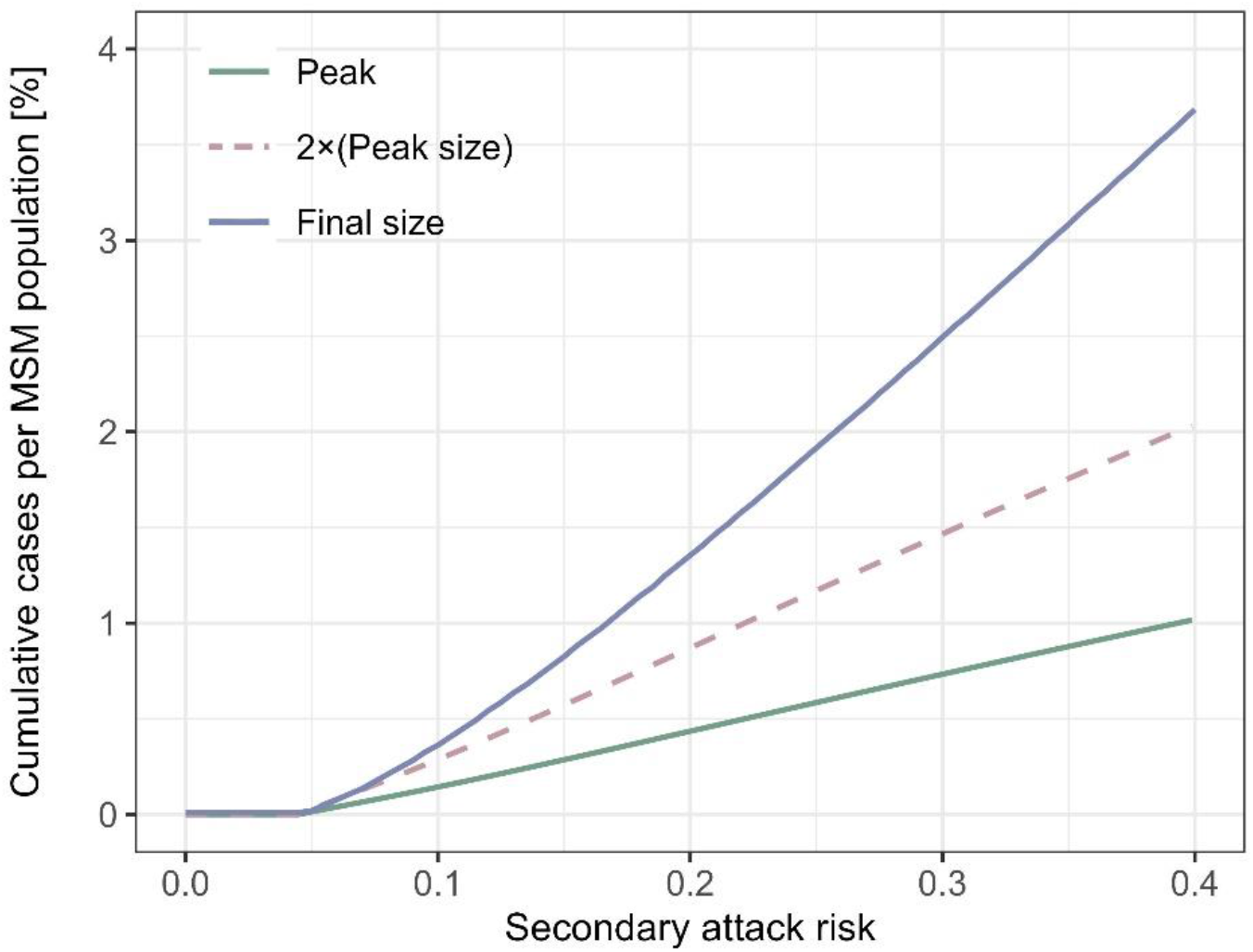
Estimated peak and final sizes by secondary attack risk in the absence of effective interventions or behavioural changes. Cumulative numbers of cases per MSM population at the peak and at the end of an epidemic estimated by our model accounting for heavy-tailed sexual partnership distribution and infection-derived immunity are shown. For comparison, a dotted line representing the double the outbreak size at the peak is also included.

Our results suggest that early infection of individuals with highest risks in a heavy-tailed sexual partnership distribution may have been sufficient to cause downward trends in monkeypox epidemics even without effective control measures. Empirical peak size data in many countries and US states with CIPPs of around 0.1–0.7% broadly corresponds to the estimated herd immunity thresholds in our model with an SAR of 10–30%. This range of SAR per sexual encounter would be plausible as it is comparable to existing estimates of non-sexually associated household SAR (*7*). However, this assumes that most infections were reported and reflected on the observed CIPPs; a higher SAR is expected (which is also possible given the close nature of sexual contacts) if cases were significantly underreported. It has been shown that the herd immunity threshold in a heterogeneous population becomes lower than the classical formulation 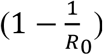, which assumes homogeneity. Britton et al. (*25*) showed in their illustrative example that introducing heterogeneity into a SARS-CoV-2 transmission model lowers the herd immunity threshold from 67% to 50%. However, compared with their example in the context of respiratory infections, heterogeneity relevant to the transmission dynamics of monkeypox is extreme due to the heavy-tailed nature of sexual contact patterns. As a result, our model replicated monkeypox epidemics over an MSM sexual contact network starting to decline even before 1% of MSM population experiences infection despite having an *R*_0_ of above 1. These epidemic dynamics driven by a highly heterogeneous sexual contact network should be accounted for when evaluating trends in the current monkeypox outbreak.

Attributing the observed decline in cases to interventions or behavioural changes without accounting for rapid accumulation of infection-derived immunity can bring a risk of misleading policy assessment. While these factors may also have had effects, our model suggests plausible scenarios in which infection-derived immunity alone could explain the observed peak sizes. The observed CIPPs in the global outbreak in 2022 ranging around 0.1– 0.7% as opposed to the classical herd immunity threshold of > 50% for an *R*_0_ of > 2 underscore the role of heavy-tailed sexual contact networks. Our model suggests that accumulation of infection-derived immunity among high-contact individuals in those networks is likely to have played a key role in limiting peak outbreak sizes. Meanwhile, we also observed variations in CIPPs that may reflect other factors including interventions, behavioural changes and case ascertainment. Although we did not find a clear correlation between CIPPs and allocated vaccine doses in US states, we found that later epidemic onset was associated with lower CIPP (Fig. S2). This may indicate possible roles of interventions and behavioural changes because places with later epidemic onset may have had more lead time to implement these early in their outbreaks. However, interpreting these observed correlations requires caution because of possible confounding—states with more cases may be more likely to be allocated more vaccines; countries and states with more active MSM populations may have been more likely to introduce monkeypox cases in the earlier phase of the outbreak. More direct and robust evidence would be required to conclude on the effects of interventions and behavioural changes in lowering epidemic peaks. A highly heterogeneous empirical partnership distribution among MSM suggests that the current monkeypox outbreak is likely driven by individuals with highest numbers of partners (*5*). Whether interventions or behavioural changes have a substantial effect on disease spread depends on their acceptance and effectiveness among those individuals. Vaccination campaigns for MSM in many settings targeted toward those with more partners (either directly assessed or using proxy measures of risk such as pre-exposure prophylaxis use and recent diagnosis of bacterial sexually transmitted infections (*26, 27*)). However, vaccine coverage data stratified by sexual behaviours has not been made widely available and the impact of this intervention is thus difficult to quantify. A voluntary reduction in sexual contacts has been reported in a recent questionnaire data from MSM in the US (*11*). Similarly, this data is not sufficient to estimate the impact on transmission dynamics as it lacked quantitative measures (respondents only answered whether they reduced their sexual contacts but not to what extent). Moreover, it is unclear whether the results are representative for individuals with the highest numbers of partners, among whom changes to partnership patterns would be most impactful on transmission. Our simulations suggest that accumulation of infection-derived immunity can plausibly reproduce the observed decline in monkeypox cases. More data is needed to discriminate the role of interventions and behavioural change from mere saturation of infection. Until this is available, attributing the decline to these factors alone may overstate their impact.

Assessing the precise contribution of interventions and behavioural changes could inform whether the current public health operations still have room for improvement. Our model projected that the declining phase of an epidemic in a heavy-tailed contact network may be gradual especially if the SAR is high. This means that, regardless of the factors driving peak incidence, promoting and providing effective and sustainable means of prevention, particularly vaccination, to those at risk—not only in newly affected countries but also in countries where monkeypox has long been endemic—remains crucial to end the global epidemic as soon as possible. Sustained resourcing is particularly important given that there might be waning of immunity or turnover in the population of MSM with the most partners, which would lead to the replenishment of susceptible individuals and therefore of epidemic potential.

Our analysis holds several limitations. MSM population size estimates used to calculate CIPP were subject to uncertainties and potential biases (*23, 24*). For example, MSM population size estimates for multiple countries are indicated to be less reliable by the authors (*23*), which may have affected some of the countries that showed outlier values of CIPP, e.g. Luxemburg. We assumed that the sexual partnership distribution in UK estimated in the previous study (*5*) applies to countries experiencing an epidemic among MSM. Some deviations from the UK partnership distributions are expected in different settings, although our sensitivity analysis suggested the robustness of our qualitative conclusions (Fig. S3). We found possible weak positive correlations between CIPPs and MSM population sizes (Spearman’s correlation 0.35 [-0.08, 0.69] among countries and 0.45 [0.12, 0.70] among US states) (Fig. S4). They may indicate variations in partnership distributions or other factors including case ascertainment between large and small countries/cities. We did not consider network assortativity or clustering, which may have led to overestimation of final size (*28*). We also assumed that the influence of imported cases on the local dynamics at country-and US state-levels are negligible; however, this may not have been the case especially in populations with a small case count. Finally, we reiterate that our findings should not be viewed as evidence on the effects of interventions and behavioural changes in the current outbreak. Our model plausibly explained CIPPs at both country-or US state-levels that are at similar order and substantially lower than the classical herd immunity threshold without needing the effect of interventions or behavioural changes. However, such patterns could also be observed if included countries and US states exhibited similar levels of interventions or behavioural changes at time of their epidemic peaks. Further studies incorporating our findings on the saturation effect from infection-derived immunity will enable us to better understand the evolving situations of monkeypox epidemiology.

## Materials and Methods

### Data source

We used the cumulative incidence data of reported monkeypox cases in countries and US states that have ever observed at least 10 cases over the successive 5 days as of 15 October 2022 (*29, 30*). Due to the data availability of MSM population size (studies where methods were systematically applied to countries and states), we limited our analysis of CIPP to European countries, Canada, all US and US states. As the cumulative case counts in US states are continuously updated on the CDC website, we referred to a public repository and an internet archive for historical data from CDC as necessary (*31, 32*). The total and estimated MSM population sizes in included countries and US states were collected from previous studies and public data sources (*23, 24, 33, 34*). The MSM population was defined as “the population which contributes to the HIV epidemic among MSM” in (*23*) and “the number of men who had sex with men within the past 5 years” in (*24*), respectively, and we assumed that they approximately represent sexually active MSM population considered in our model. We also used the data of allocated doses of vaccine in US states as of 15 September 2022 (*12*). Similar data on vaccination across countries was not available.

### Estimation of peak size

To identify the period when an epidemic was most likely reaching its peak, we fitted a Gompertz curve to cumulative incidence in each of the included countries and US states, which serves as an approximation of saturating epidemic growth (*35, 36*). Let *F*(t) be the Gompertz function:

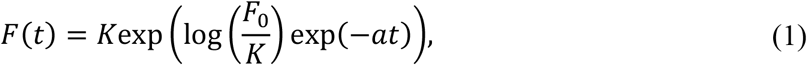

where a (decay rate of the growth), *F*_*0*_ (initial number of cases) and *K* (carrying capacity) are the parameters that characterise the Gompertz function.

We defined the local growth rate of the Gompertz curve at time *t* as:

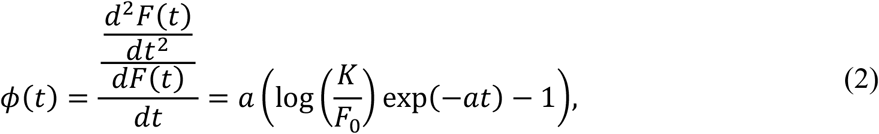

Note that, because 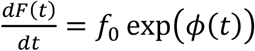 satisfy the condition 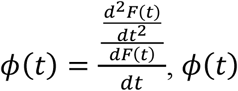 can be interpreted as the growth rate of an exponential function that locally approximates the epidemic curve. The peak of an epidemic is characterised as the point where the local growth rate (ϕ) is zero: around this point the cumulative incidence is expected to be approximately linear. We therefore considered the period during which the estimated ϕ is sufficiently close to zero (|ϕ| ≤ *0*.*0*1) as the possible time range for the epidemic peak. We assumed that the observed cumulative incidence follows the Gompertz function with a normally-distributed errors, i.e. Normal(mean = *F*(t), sd = σ). We estimated a, *F*_*0*_, *K*, and σ using the Markov-chain Monte Carlo (MCMC) method (via the {rstan} package in R (*37*)). We employed an improper flat prior (Uniform(*0*, ∞)) for σ and weakly-informative priors for a, *F*_*0*_ and *K*: HalfNormal(*0*, 1) for a (given that it would below 1), LeftTruncatedNormal(1*0*, 5*0*) for *F*_*0*_ truncated at 0 (reflecting that all the included countries/US states hold at least 10 cases) and HalfNormal(*0*, *0*.*0*25*N*_*c*_) for *K*, where *N*_*c*_ is the total population size of country *c*. The prior for *K* reflects that carrying capacity would be at least under 2.5% of the total population size, given that the relative MSM population size is reported to be up to 5.6% of the adult male population size in Europe. We obtained 10,000 MCMC samples from four chains using the Hamiltonian Monte Carlo algorithm with No-U-Turn-Sampler, where the first 2,000 warm-up iterations were discarded. The resulting MCMC samples showed an R-hat statistic of below

1.05 and an effective sample size of at least 200. We identified the period where the 95% credible interval of estimated local growth rate contains a value within the range |ϕ| ≤ *0*.*0*1 and defined CIPP as the cumulative number of cases per MSM population size during this period (the upper limit of CIPP may be undefined if the epidemic has not yet clearly passed the peak). We then constructed a consensus range of CIPP across countries (“global consensus range”) and US states (“US consensus range”) included in the analysis. We defined the consensus range as a set of values that lie within the CIPP of at least 50% of included countries/states.

### Epidemic model

To describe the heterogeneity of sexual contact networks among men who have sex with men (MSM), we used a left-truncated Weibull distribution, *w*(*x*), estimated to represent the number of sexual partners over the infectious period of monkeypox elsewhere (*5*). In the original study, the distribution was fitted to the annual sexual partnership data for MSM in the UK and then rescaled to the assumed infectious period of monkeypox of 21 days. In the present study we instead assumed 14 days to reflect updated epidemiological knowledge (*38, 39*). The resulting Weibull distribution of the mean number of partners over 14 days has a shape parameter *α* = 0.10 and a scale parameter *θ* = 5.2×10^−11^. Alternatively, the distribution is characterised by a Pareto-approximated exponent κ = 1.1 (i.e. the exponent parameter of a Pareto distribution that approximates the body part of the Weibull distribution (*5*)) and the upper 1 percentile of 16.

We developed a dynamical model of monkeypox transmission that accounts for the heavy-tailed sexual partnership distribution among MSM. Instead of calendar time, we use the cumulative force of infection (CFOI) to measure progression of an epidemic for mathematical convenience. Let *S*(Λ, *x*) be the population fraction of susceptible individuals with degree x when CFOI is Λ. We assumed that the population is fully susceptible at the start of an epidemic, i.e. 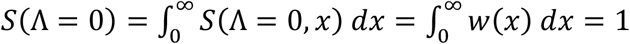. We assumed that individuals are exposed to monkeypox virus at a probability proportional to their degree (i.e. those with a large number of partners are more likely to be chosen) and that infected individuals develop permanent immunity to be protected from future infections while retaining their original sexual behaviour for the rest of the epidemic. The process of depletion of susceptibles is then described as

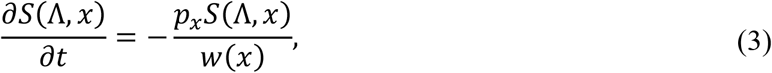

where *p*_*x*_ is the relative frequency of *x* among cases 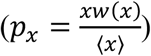 and ⟨*x*⟩ is the mean degree 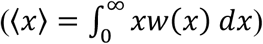. Solving Equation (3), we get a closed form of *S*(Λ, *x*) and cumulative incidence proportion *I*(Λ) as:

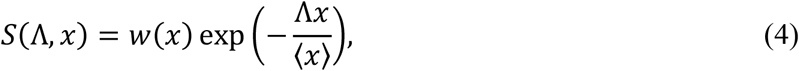

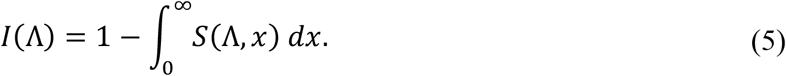

The effective reproduction number (*R*_eff_) over a sexual contact network is then derived using the mean excess degree of susceptibles. The mean excess degree of susceptibles (⟨*e*_*S*_(Λ)⟩) is given as:

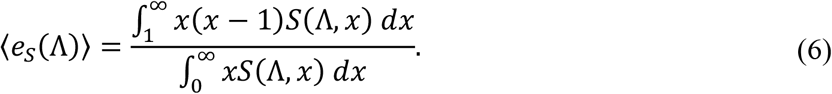

Denoting the secondary attack risk (SAR) per sexual partnership as *β*, we obtain the following equation for *R*_eff_:

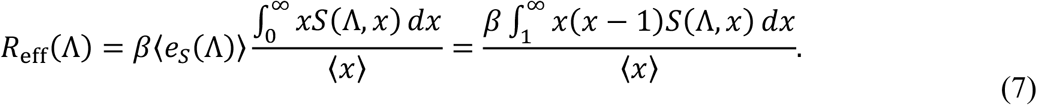

Note that when Λ = *0*, i.e. at the start of an epidemic, *R*_eff_(Λ) corresponds to the basic reproduction number (*R*_0_) as defined in (*5*).

The peak of an epidemic is the point where *R*_eff_(Λ) = 1. By rearranging Equation (7), we get an estimator for SAR (β):

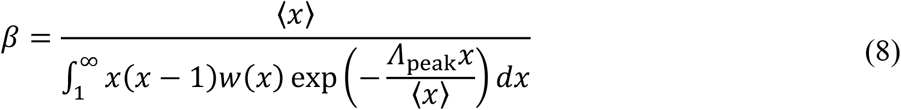

where Λ_peak_ is the CFOI at the peak of an epidemic. We estimated SAR that renders our model consistent with the global and US consensus ranges of CIPP using Equations (5) and (8); that is, assuming that interventions or behavioural changes have negligible effects on peak sizes and that case ascertainment was sufficiently high, the estimated consensus range of CIPPs corresponds to *I*(Λ_peak_), which allows us to compute *β*. We assumed that observed cases are predominantly among MSM who acquired infection via sexually-associated contacts (*3, 5*) and that the transmission dynamics is thus well described by spread within a closed MSM population.

We also derived the expected final size of an epidemic using the following equation on the CFOI at the final size (Λ_final_):

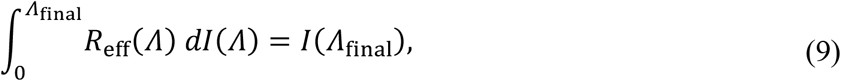

All the analysis was conducted either in R v. 4.0.2 or Julia v.1.7.2. Replication codes are available on a GitHub repository: (https://github.com/hiroaki-murayama/MPX_depletion_susceptibles)

## Data Availability

All data used in this study are publicly available

## Acknowledgements

AE and FM are supported by Japan Society for the Promotion of Science (JSPS) *KAKENHI* (to AE, grant number: 22K17329 and to FM, 20J00793), JSPS Overseas Research Fellowships (to AE). AE and HM are supported by foundation for the Fusion Of Science and Technology (to AE). SF and SA are supported by Wellcome Trust (to SF, 210758/Z/18/Z). CABP is supported by the Innovative Medicines Initiative 2 Joint Undertaking under grant agreement EBOVAC3 (800176); this Joint Undertaking receives support from the European Union’s Horizon 2020 research and innovation programme and EFPIA. S-mJ is supported by the Centers for Disease Control and Prevention (CDC) SHEPheRD (200-2016-91781). EF is supported by the Medical Research Council, United Kingdom Research and Innovation (MRC-UKRI), MR/S020462/1.

## Author contributions

Conceptualization: A.E. Methodology: A.E., H.M., C.A.B.P. Investigation: A.E., H.M., Visualisation: H.M. Funding acquisition: A.E., S.F., C.A.B.P., E.F. Writing–original draft: A.E., H.M. Writing–review and editing: C.A.B.P, S.A., F.M., S-m.J., E.F., S.F.

## Supplementary Materials

### Relationship between peak and final sizes in a homogeneously mixing population

We showed in Fig. 2 that the final size of an epidemic over a heavy-tailed sexual contact network in our model is generally larger than double the size of the peak. Here we provide a quick proof that this is contrary to the feature of a homogeneous mixing Susceptible-Infectious-Recovered (SIR) model. Let *y* and *z* represent the cumulative incidence per capita at the peak (peak size) and at the end of an epidemic (final size), respectively. These are defined by the following equations in a homogeneous mixing SIR model (*40*):

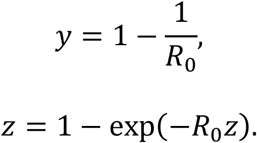

The ratio between the peak and final sizes is then given as 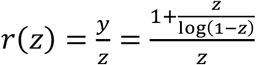. We can show that *r*(*z*) is a monotonically increasing function for *0* < *z* < 1 as follows. The derivative of *r*(*z*) is

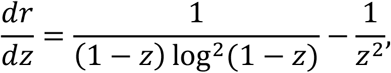

whose sign matches that of *z*^2^ − (1 − *z*) log^2^(1 − *z*). We get log(1 − *z*) < −*z* < *0*, which yields

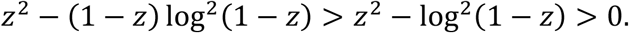

With 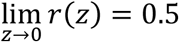, it is assured that *r* > 0.5, i.e. the final size of an epidemic is always smaller than double the peak size.

## Additional Figures

**Fig. S1.**
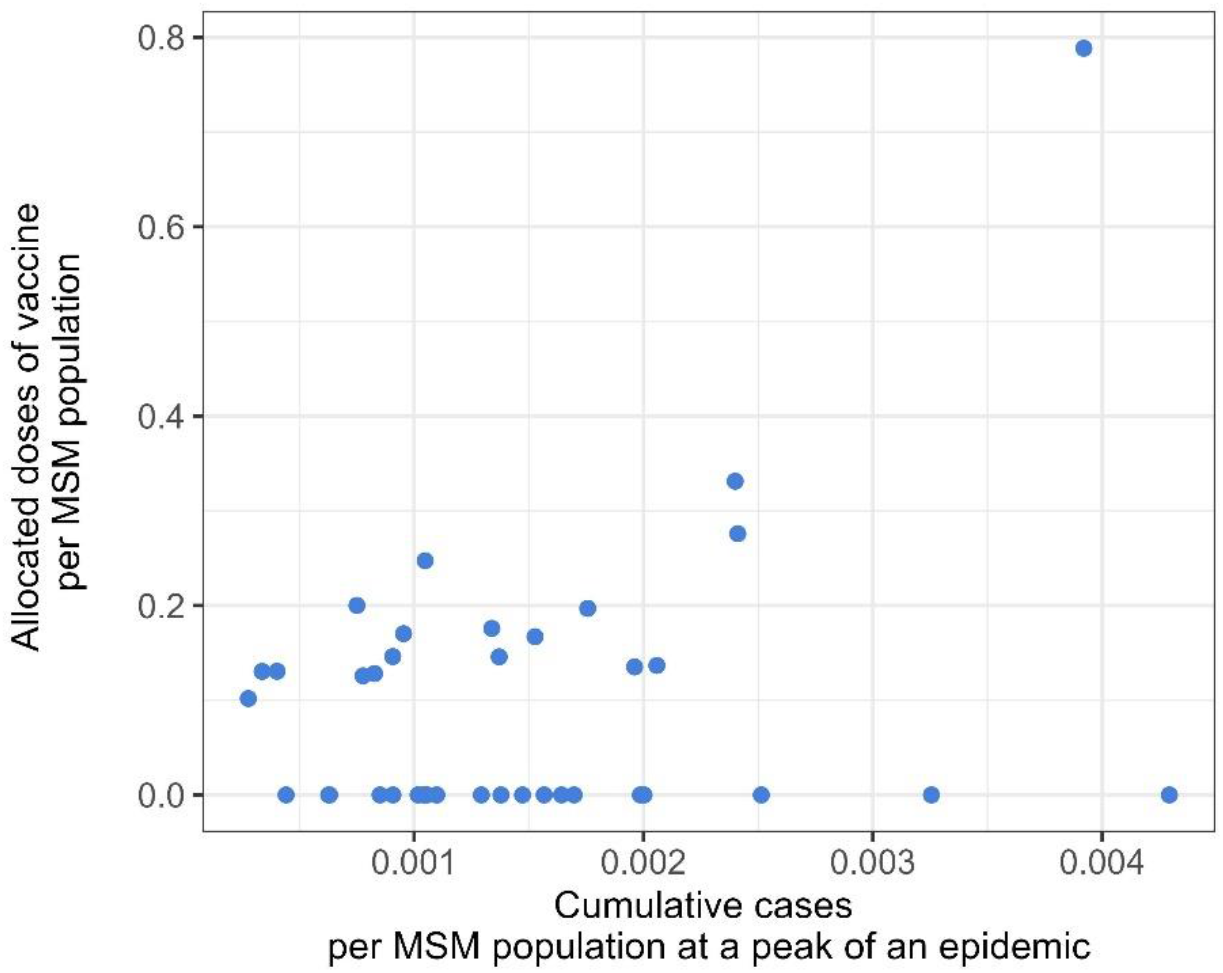
Correlation between CIPP and allocated doses of vaccine by the peak among US states. The lower limits of CIPP in US states are displayed as dots. The Spearman’s correlation coefficient is 0.21 [95% confidence interval: -0.12, 0.53].

**Fig. S2.**
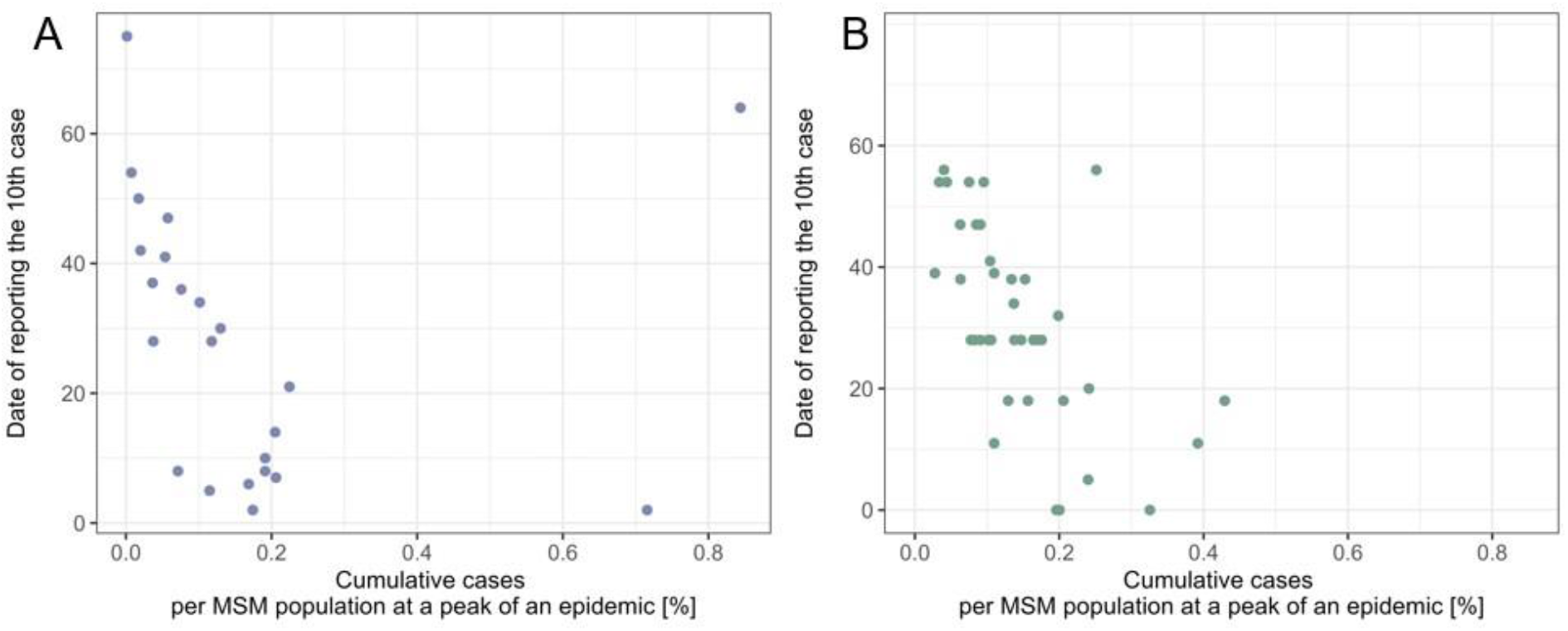
Correlation between CIPP and date of reporting the 10th case. The lower limits of CIPP (A) by country and (B) by US-state are displayed as dots. The Spearman’s correlation coefficients are -0.60 [-0.91, 0.11] (country) and -0.65 [-0.84, -0.37] (US states).

**Fig. S3.**
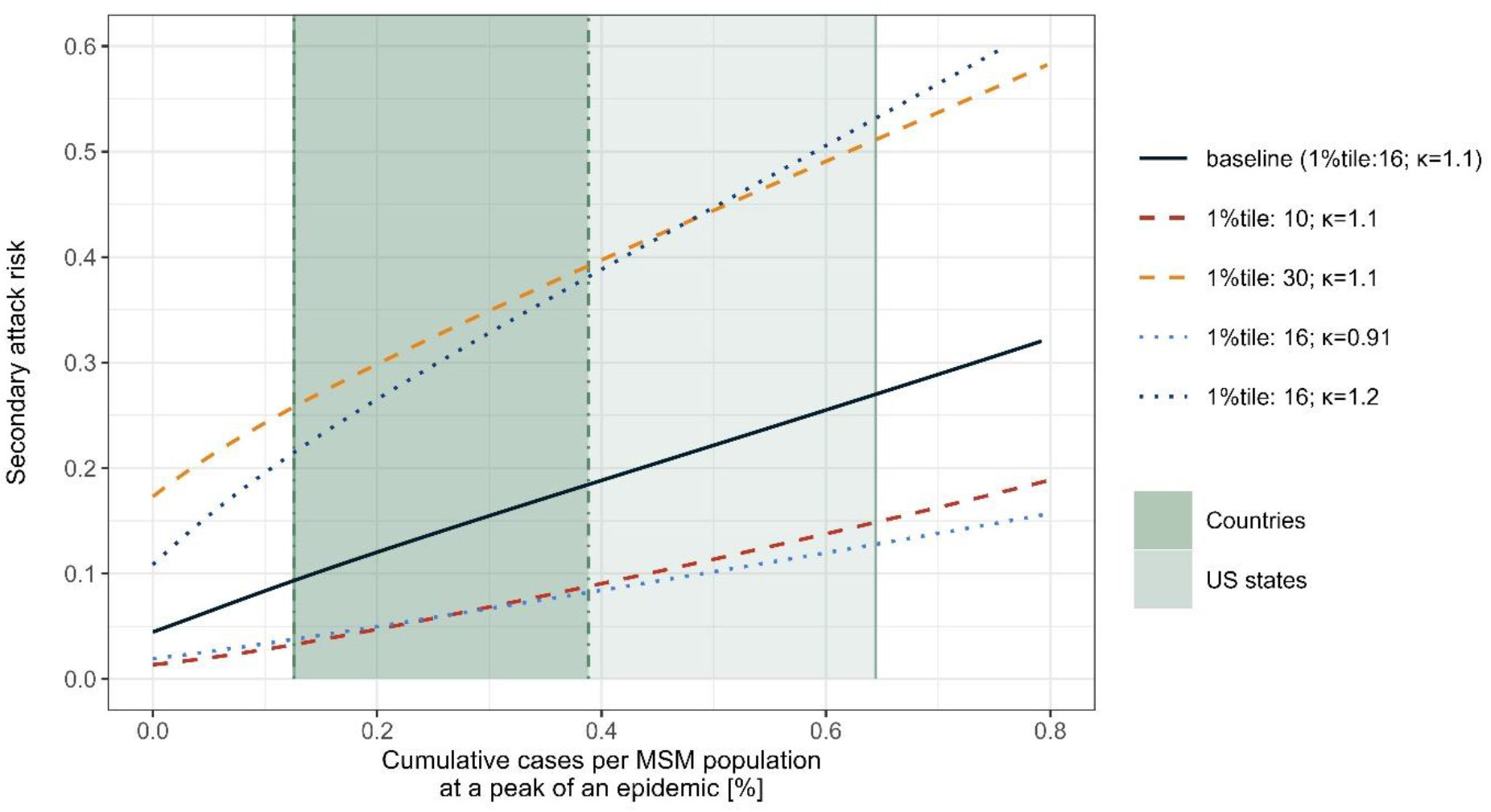
Sensitivity to the variations in the MSM sexual partnership distribution. Each dotted line shows the relationship between CIPP and secondary attack risk reflecting possible variations in the Weibull distribution representing the sexual partnerships among MSM. Changes to the assumed Weibull distribution was represented by a Pareto-approximated exponent *κ* and the upper 1st percentile. Thick and thin green areas represent the global and US consensus ranges of CIPP, respectively.

**Fig. S4.**
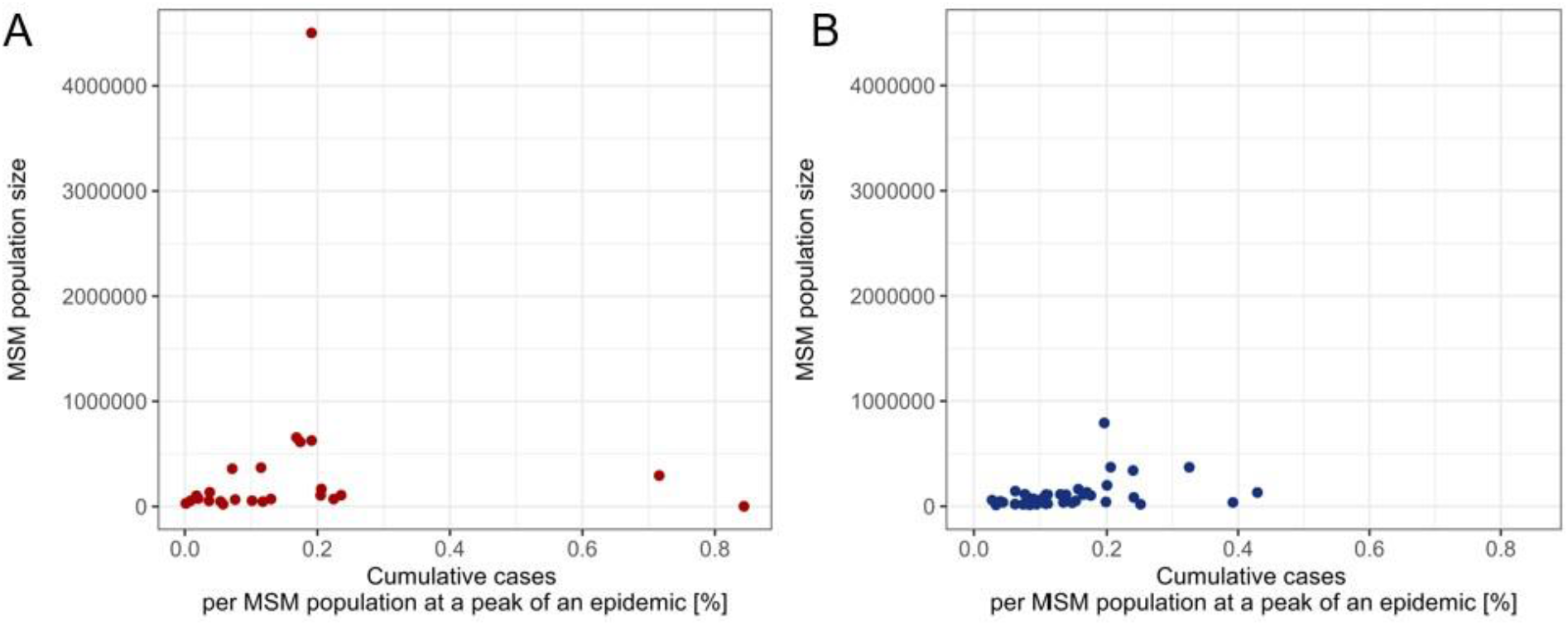
Correlation between CIPP and MSM population size. The lower limits of CIPP (A) by country and (B) by US-state are displayed as dots. The Spearman’s correlation coefficients are 0.35 [-0.08, 0.69] (country) and 0.45 [0.12, 0.70] (US states).

**Fig. S5.**
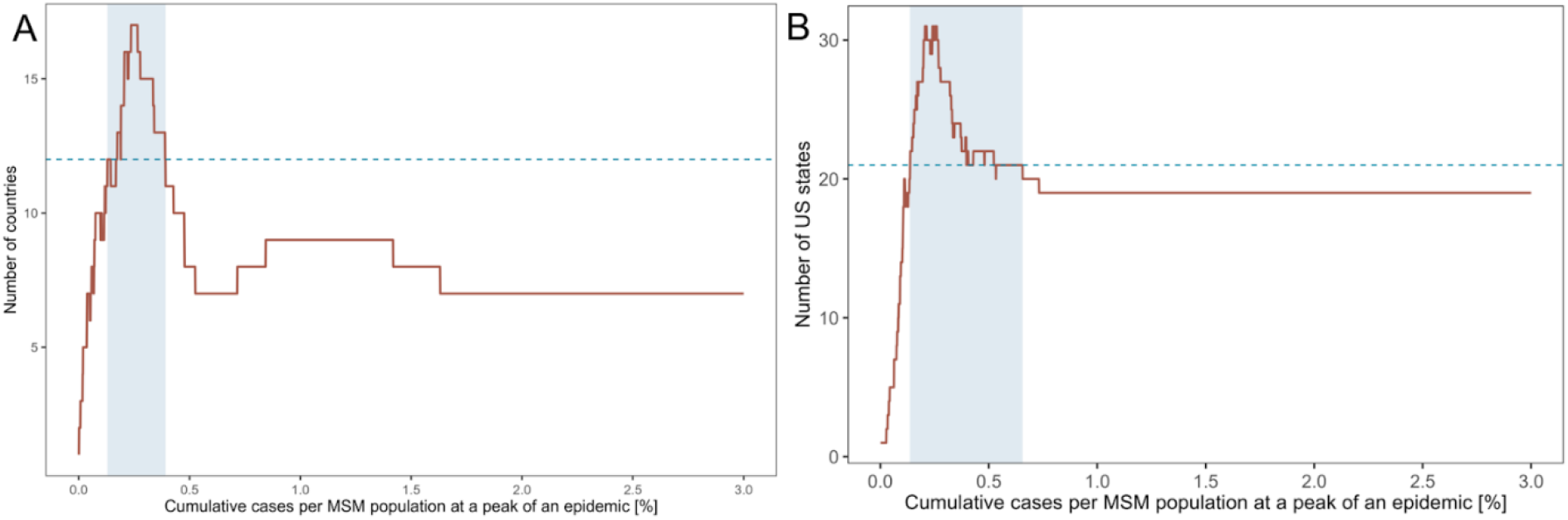
Consensus ranges of CIPP. Red line shows the number of countries/states whose CIPP includes the specific CIPP value. The light blue shaded areas represent the consensus ranges of CIPP. Blue dot line shows the threshold of 50% used to define the consensus ranges for (A) included countries and (B) US states.

**Fig. S6.**
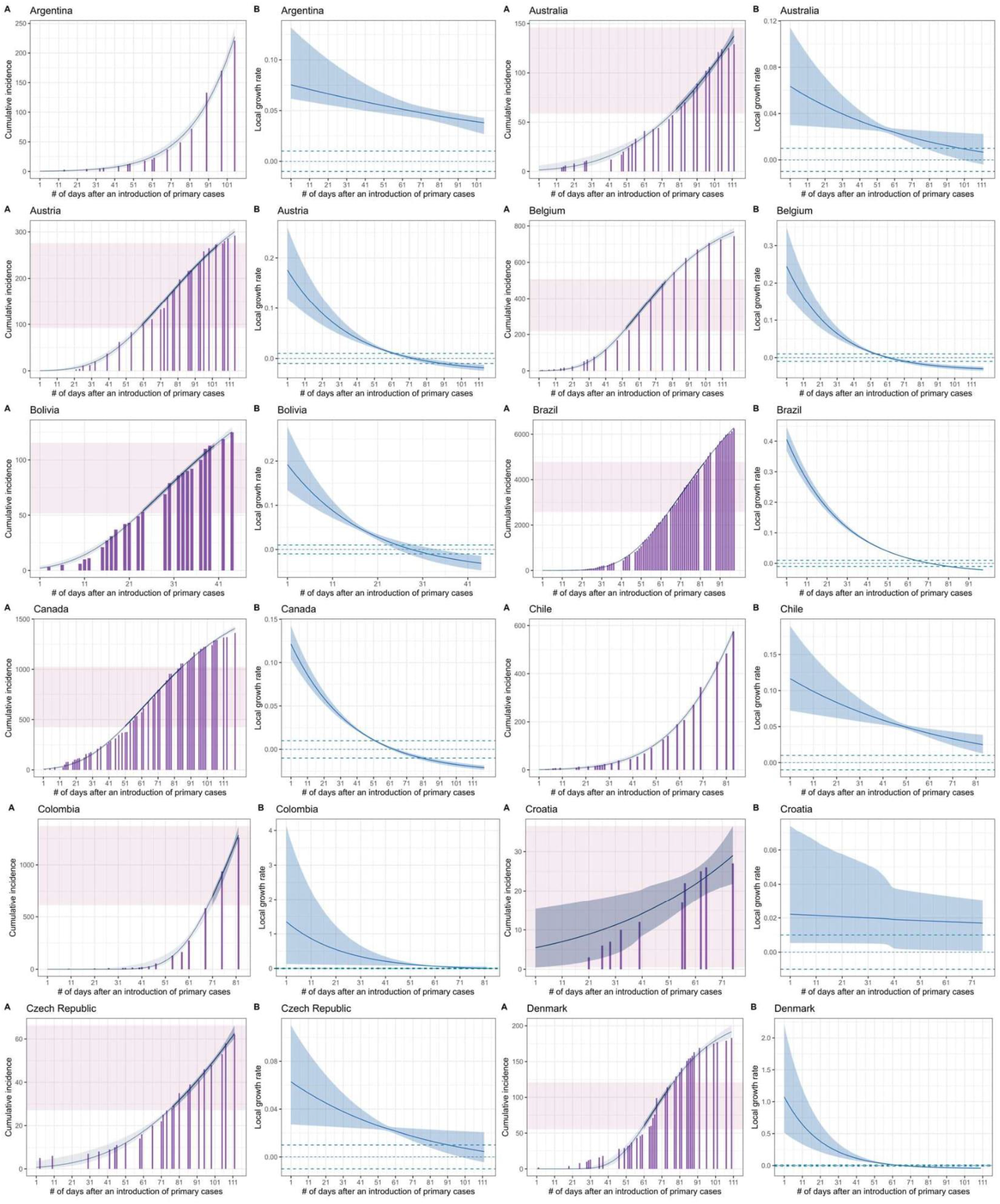
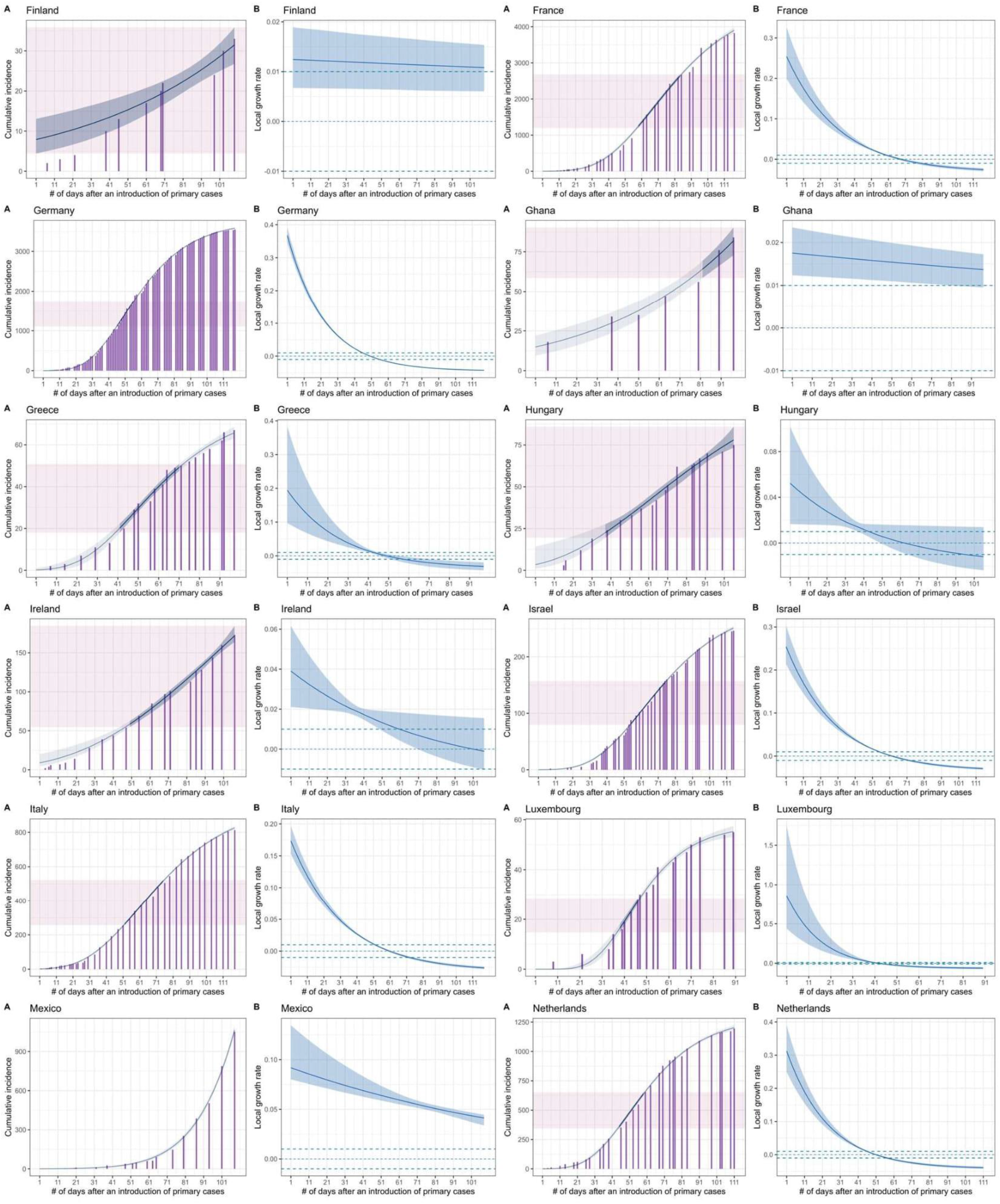
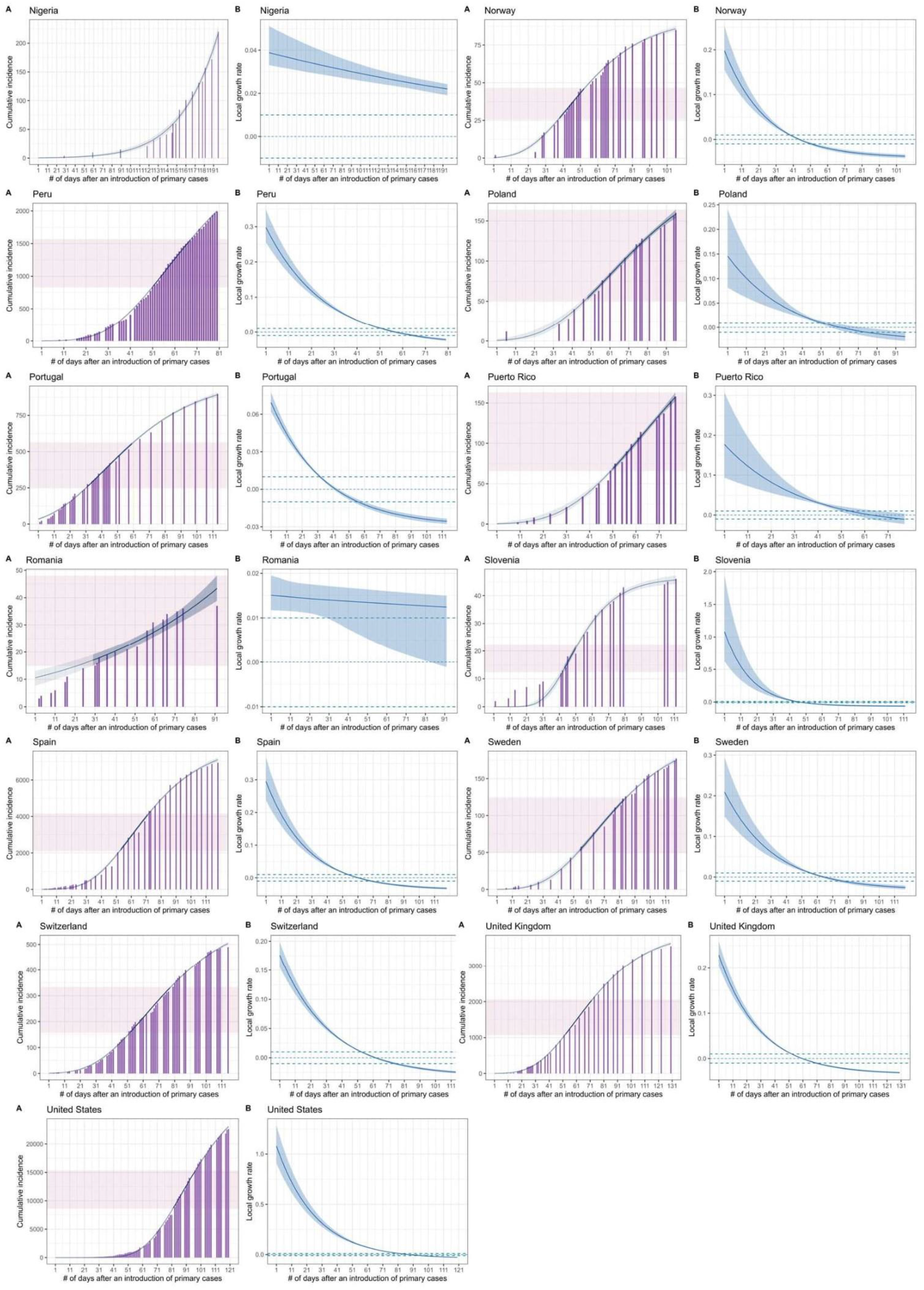
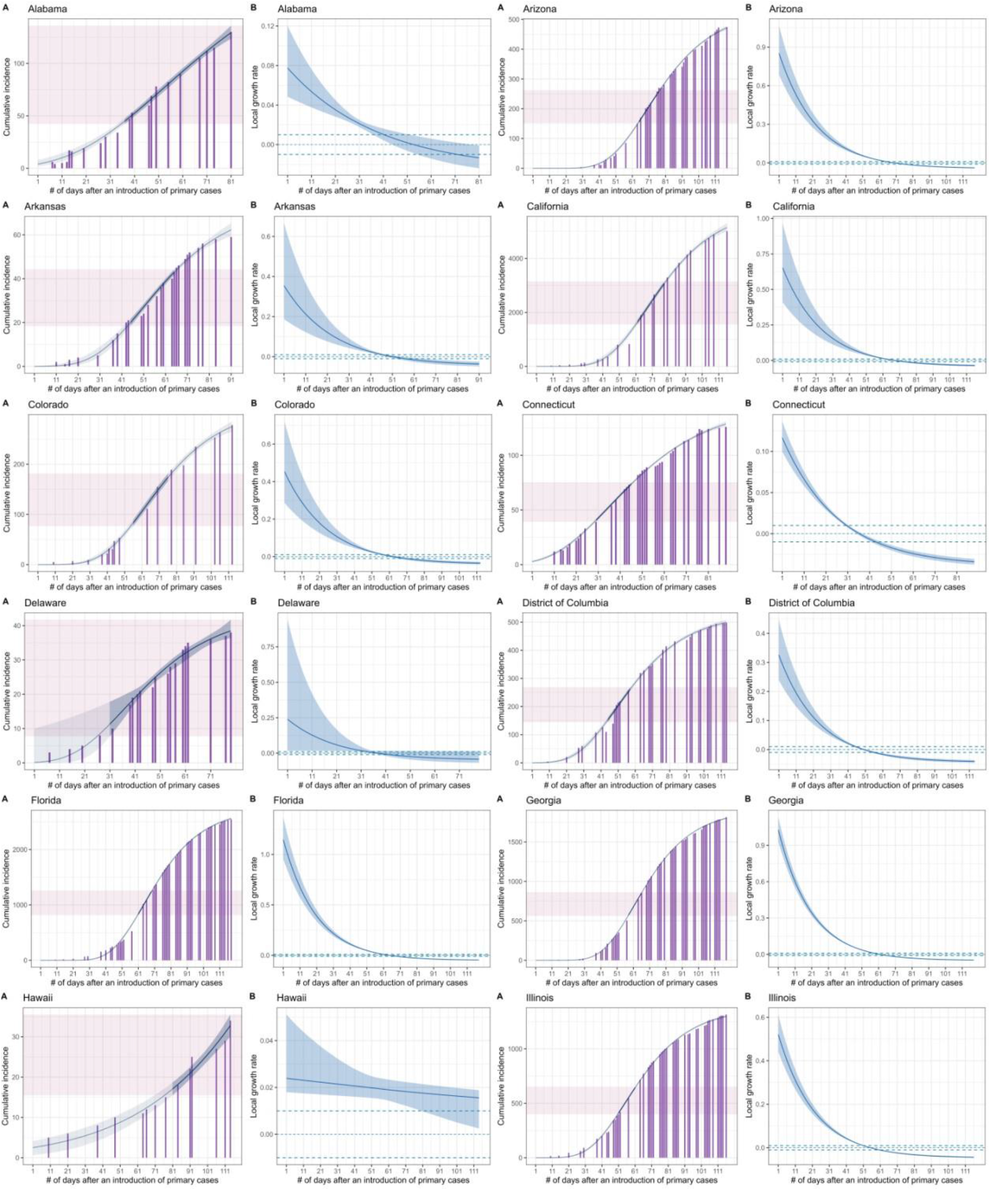
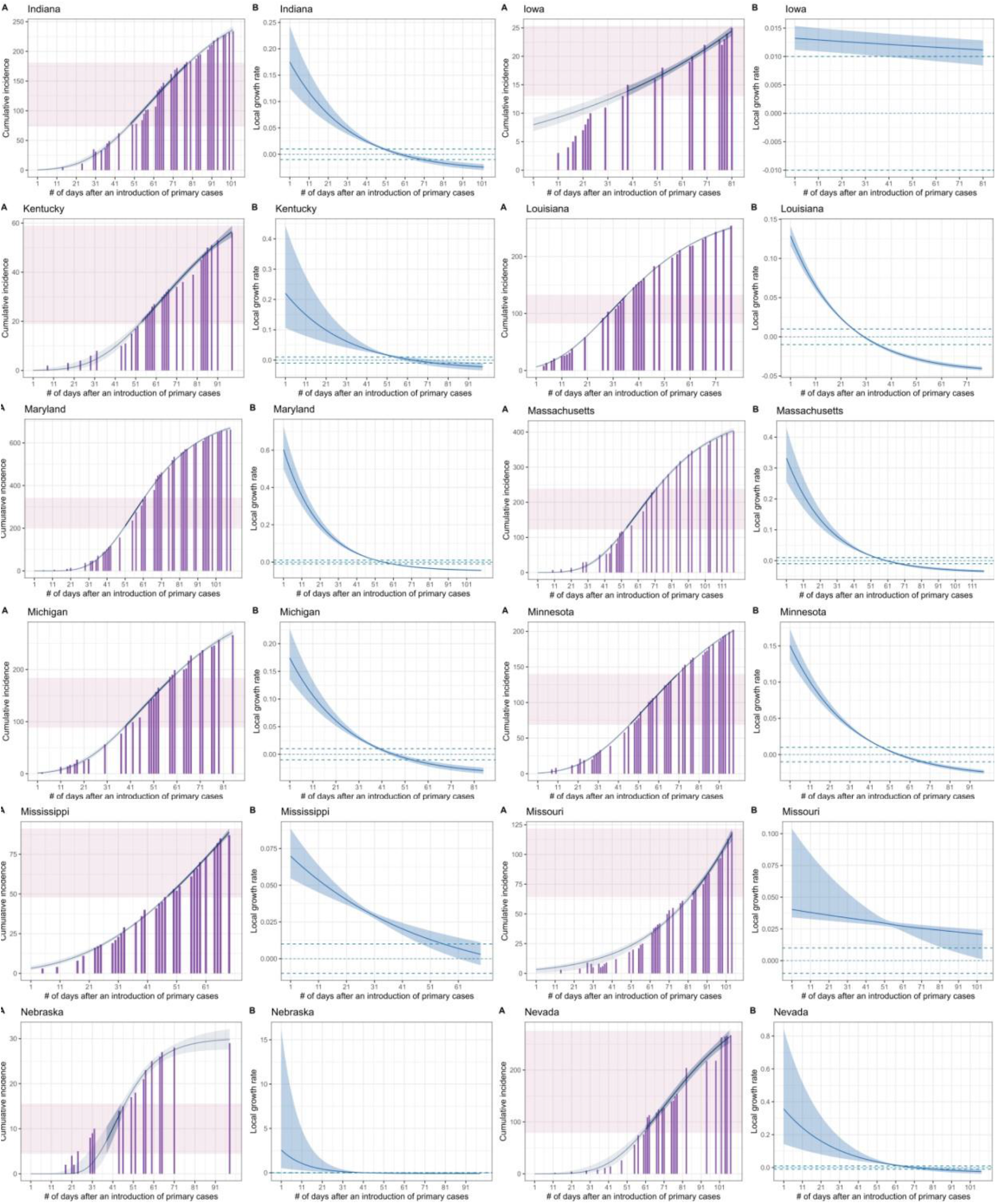
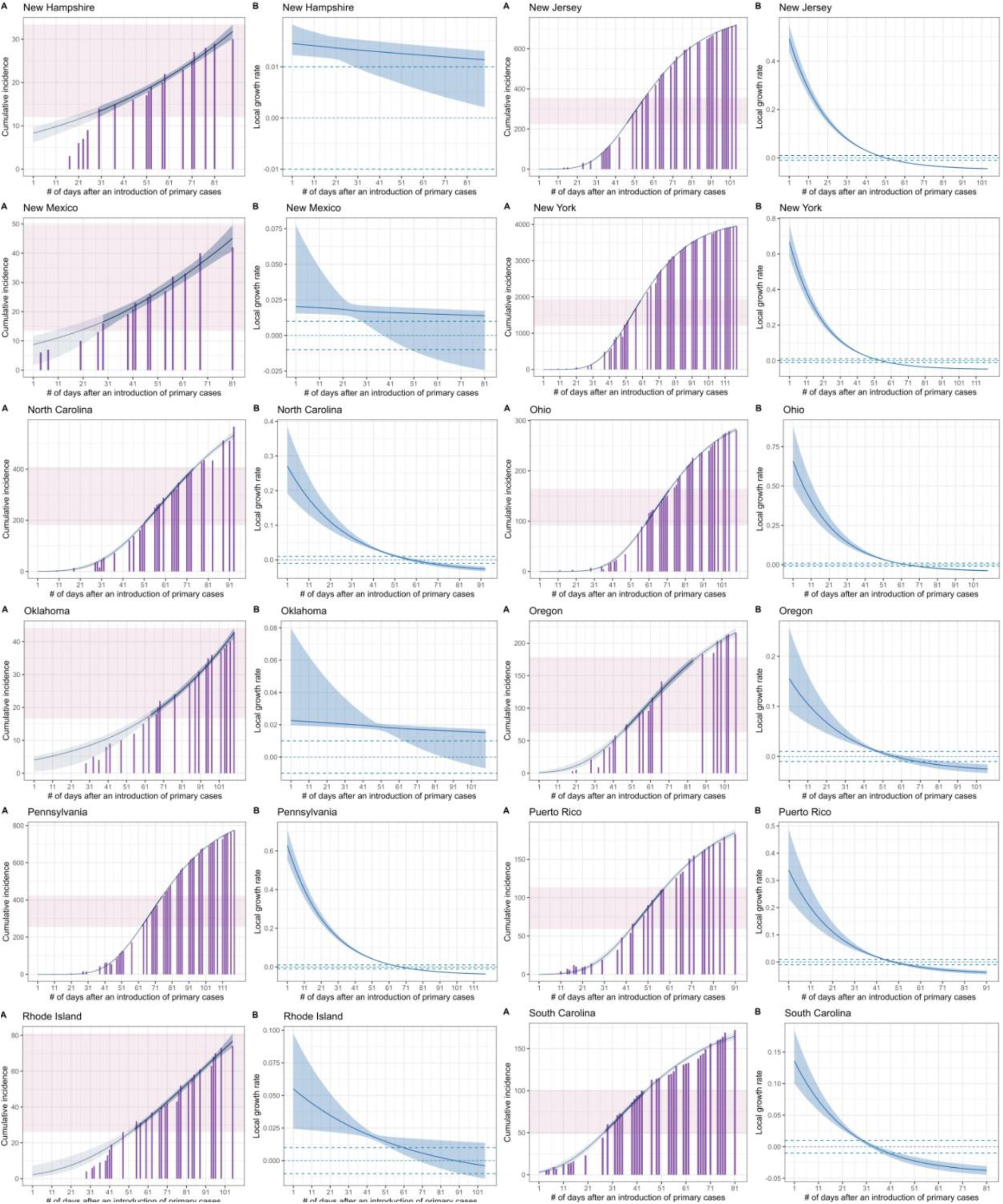
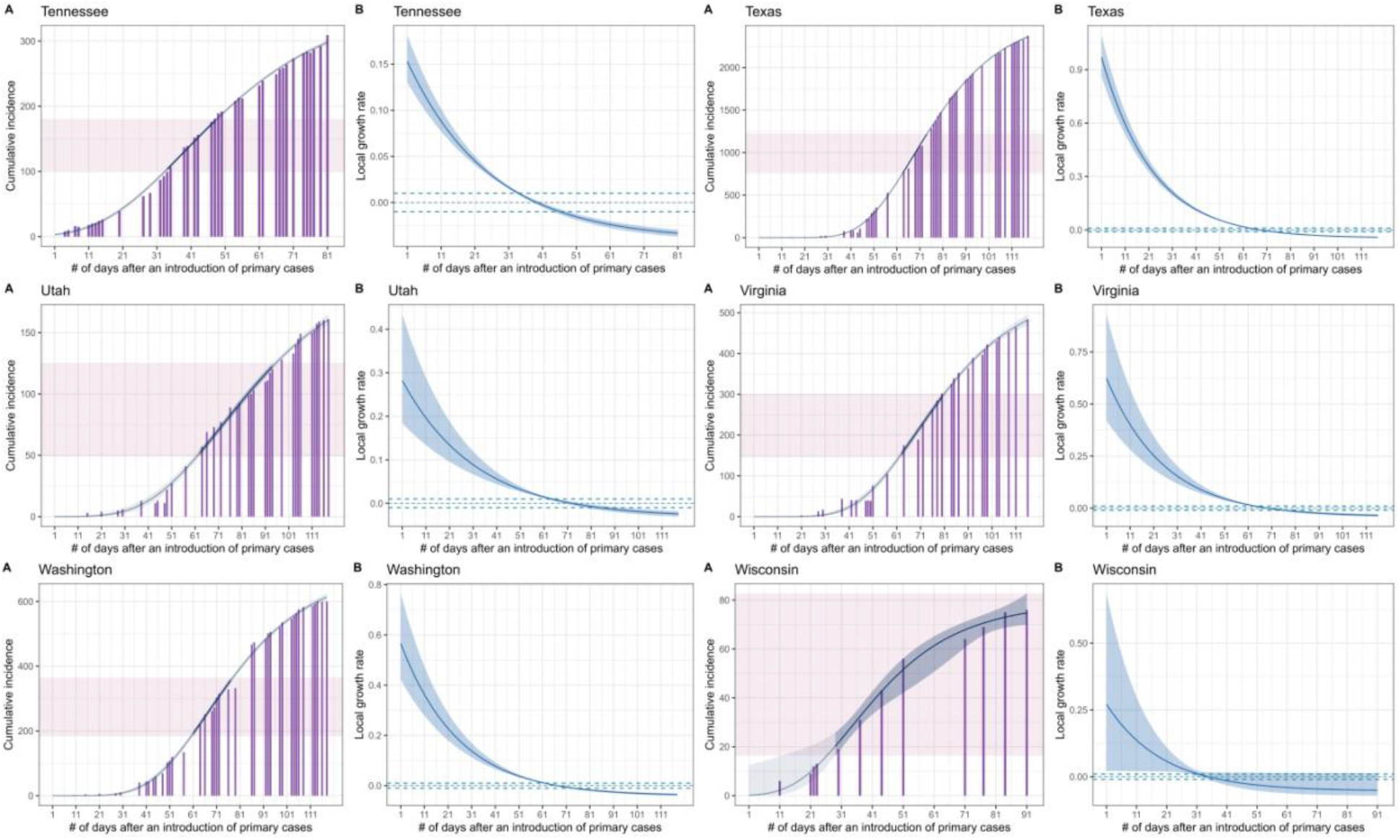
Fitted Gompertz curves and estimated local growth rates in countries and US states. Purple bars represent the reported cumulative number of monkeypox cases. Thick blue lines show the median estimates of (A) epidemic curves and (B) growth rates and blue shaded areas their 95% credible intervals. Thin purple areas show the range where the growth rate takes a near-zero value (i.e. within ±0.01).

